## Supplementary files for "Do lifestyle factors affect Patient-RepOrted clinical outcomes after total Knee replacement surgery? A feasibility cohort study (PRO-Knee)"

**Supporting Information**

**S1 Appendix.**

**Management of Patient reported outcomes data.**

| **Variable and measurement tool** | **Scoring** | **Missing data** |
| --- | --- | --- |
| Awareness of operated knee  Forgotten Joint Score | 12 questions providing an overall score between 0-100 | Outcome measure withdrawn if >3 missing values. |
| Knee pain and function  Oxford Knee Score | 12 questions providing an overall score between 0-48 | Sub-scale mean to be used if <3 missing values.  Outcome measure withdrawn if >2 missing values. |
| Health Related Quality of Life  EQ5D5L Index | All values were indexed. −0.594 (worst state) to 1.0 (best state). | Multiple imputation to be used for missing values. |
| Anxiety  Hospital Anxiety and Depression Scale (HADS) | 0-21  Those with score >10 will be compared to those <11 | Sub-scale mean used for missing values providing > 50% of answers provided. |
| Depression  Hospital Anxiety and Depression Scale (HADS) | 0-21  Those with score >10 will be compared to those <11 | Sub-scale mean used providing that more 50% of answers in total were provided |

**S2 Appendix.**

**Number of lifestyle factors and clinical outcomes for all participants**

|  | **Forgotten Joint Score** | | | **Oxford Knee score** | | |
| --- | --- | --- | --- | --- | --- | --- |
| **No of lifestyle factors** | **Baseline** | **3-months** | **6-months** | **Baseline** | **3-months** | **6-months** |
| **1 Lifestyle factor**  No. of participants  Mean score  Standard deviation | 13  **87.5**  7.8 | 13  **64.6**  31.0 | 11  **61.1**  26.1 | 13  **20.5**  7.5 | 12  **29.7**  11.0 | 11  **33.9**  7.3 |
| **2 Lifestyle factors**  No. of participants  Mean score  Standard deviation | 21  **91.5**  8.3 | 18  **78.1**  19.4 | 17  **78.5**  18.6 | 21  **16.9**  7.4 | 18  **25.2**  8.5 | 17  **26.3**  9.5 |
| **3 Lifestyle factors**  No. of participants  Mean score  Standard deviation | 6  **87.2**  12.2 | 6  **77.9**  27.3 | 6  **61.0**  36.6 | 6  **18.7**  8.0 | 6  **27.8**  12.2 | 6  **30.5**  13.9 |

**S3 Appendix**

**Satisfaction with aspects of outcome at 6-months post total knee replacement**

| **How satisfied are you with…** | **Very dissatisfied** | **Dissatisfied** | **Neither satisfied nor dissatisfied** | **Satisfied** | **Very satisfied** | **Missing data** |
| --- | --- | --- | --- | --- | --- | --- |
| **the changes you have experienced in your pain due to your knee replacement?** | **2/35**  **5.7%** | **3/35**  **8.6%** | **7/35**  **20.0%** | **10/35**  **28.6%** | **13/35**  **37.1%** | **5/40**  **12.5%** |
| **the changes you have experienced in your walking due to your knee replacement?** | **2/35**  **5.7%** | **4/35**  **11.4%** | **9/35**  **25.6%** | **7/35**  **20.0%** | **13/35**  **37.1%** | **5/40**  **12.5%** |
| **the appearance of your knee replacement?** | **1/35**  **2.8%** | **3/35**  **8.6%** | **2/35**  **5.7%** | **15/35**  **42.9%** | **14/35**  **40%** | **5/40**  **12.5%** |
| **the changes you have experienced in your ability to take part in hobbies / leisure activities** | **3/34**  **8.8%** | **3/34**  **8.8%** | **13/34**  **38.2%** | **6/34**  **17.6%** | **9/34**  **26.4%** | **6/40**  **15.0%** |
| **the outcome of your knee replacement at this point?** | **3/35**  **8.6%** | **5/35**  **14.3%** | **7/35**  **20.0%** | **8/35**  **22.9%** | **12/35**  **34.3%** | **5/40**  **12.5%** |
